# Bilateral Deep Brain Stimulation of the Subthalamic Nucleus or Globus Pallidus Internus Improves Gait Impairment in Parkinson’s Disease

**DOI:** 10.1101/2025.09.15.25332975

**Authors:** Jamal Al Ali, J. Lucas Mckay, Joe R. Nocera, Faical Isbaine, Julie T. Tran, Paola Testini, Shirley D. Triche, Christine D. Esper, Pratibha Aia, Laura M. Scorr, Lenora Higginbotham, Richa Tripathi, Nicholas Au Yong, Svjetlana Miocinovic, Cathrin Buetefisch

## Abstract

**Background and objectives:** The effect of deep brain stimulation (DBS) on gait in patients with Parkinson’s disease (PD) is variable but prospective, long-term studies—especially including globus pallidus interna (GPi) stimulation—remain sparse. We tested the hypothesis that subthalamic nucleus (STN) and GPi DBS exert acute and chronic effects on gait in patients with PD.

**Methods:** Gait kinematics were collected prospectively on patients with PD with bilateral or unilateral STN or GPi DBS, at baseline before initial DBS activation (n=104), acutely after activation (n=102), and chronically at 1-month (n=75) and 12-months (n=82). Gait speed was the main outcome measure. Kinematic measures of pace, rhythm, and variability and clinical scales were secondary outcome measures.

**Results:** The average gait speed at baseline in levodopa off state was abnormally slow (80.6 cm/s). DBS activation acutely increased gait speed (97.4 cm/s, p < 0.001) which was maintained at 1- (93.0 cm/s, p < 0.001) and 12-month follow up (91.2 cm/s, p < 0.001). Acute changes during initial programming predicted chronic gait outcomes. When DBS targets were analyzed separately, only patients receiving bilateral STN or GPi DBS improved gait speed along with kinematic measures of pace, rhythm, and variability. Preoperative levodopa response of the MDS-UPDRS axial sub-score correlated with gait speed response to DBS while the total score did not.

**Discussion:** Patients who received either bilateral STN or GPi DBS improved gait kinematic in levodopa off state at 1 year so both targets may be considered for treatment of gait dysfunction, unlike unilateral implantations which resulted in no change. Acute effects of STN or GPi DBS on gait speed should be considered when programming patients as they predict chronic outcomes. When determining DBS candidacy, the degree of axial symptom improvement with levodopa should be considered.

## INTRODUCTION

Subthalamic nucleus (STN) and globus pallidus internus (GPi) deep brain stimulation (DBS) have been highly effective as a treatment for appendicular symptoms (tremor, rigidity, bradykinesia) of Parkinson’s disease (PD). The effect of DBS on gait, however, is less predictable and is thought to develop over hours to days after DBS activation ^1,2^. Therefore, the management of gait abnormalities with DBS can be difficult in clinical practice ^3–5^. Only a limited number of longitudinal studies have followed patients with PD after DBS, with most concentrating on stimulation of STN ^6^, while less is known about the effects of GPi DBS on gait ^7,8^.

Barriers to the development of effective DBS programming algorithms for gait abnormalities include the limitation of clinical scales to capture DBS-related gait changes, the focus on the upper extremity motor symptoms during DBS programming, and the paucity of studies systematically reporting the effect of stimulation parameters on gait ^3^. More recently, objective kinematic approaches such as wearable sensors and instruments walking mats have been developed to improve quantification of gait abnormalities and accurately follow changes in gait over time ^9,10^. The kinematic measure of gait speed has been considered a clinical meaningful outcome measure, as it has been demonstrated to correlate with health care outcomes. Gait speed above 100 cm/s is thought to be necessary for normal functioning in the community ^11–17^, and ±10 cm/s change as a clinically meaningful difference in older adults and patients with PD ^18,19^.

In this study, we hypothesized that STN or GPi DBS exert acute and chronic effects on gait speed. The hypothesis is built on the evidence that DBS alleviates pathologic beta oscillatory activity in the basal ganglia and beta-gamma synchronization in the primary motor cortex on short timescales (seconds to minutes) ^20–22^ and that long-term effects depend on reorganization of underlying stimulated neuronal networks ^23–25^. In a prospective, longitudinal study design, we evaluated gait speed in 104 patients with PD prior to DBS activation (Baseline), immediately after DBS activation (Acute), after one month (Chronic 1-month), and after one year of stimulation (Chronic 12-month). The effects of STN or GPi DBS on additional gait kinematic measures across five gait domains (pace, variability, rhythm, postural control, and asymmetry) were explored as secondary outcomes.

## METHODS

### Study Design

This is a prospective, longitudinal cohort study in patients with PD implanted with STN or GPi DBS electrodes. Work up for DBS candidacy, DBS target selection, surgery and programming were performed according to an established clinical protocol in keeping with current guidelines ^26–28^ and were not influenced by study participation. Study data were collected during three visits. The first visit was at the initial DBS programming approximately 4 weeks after lead implantation, where data were collected before DBS activation (“Baseline”) and after 5 minutes of stimulation at clinically optimized settings (“Acute”).

Additional data were collected at the 1-month programming visit (within 20-60 days from initial programming, “Chronic 1-month”) and at the 12-month programming visit (within 9-16 months from initial programming, “Chronic 12-month”). At Chronic 1-month and Chronic 12-month, measurements were done prior to any DBS adjustments. Blinding was not possible as each time point had one pre-specified condition. Gait testing at each visit was performed in the clinically defined OFF state, approximately 12 hours after the last PD medication intake.

### Patients

All patients with PD who had first-time STN or GPi DBS surgery from June 2020 to July 2024 were assessed for eligibility (n =122). Prior to surgery the patients completed extensive clinical, neuropsychological, psychiatric and motion testing to meet clinical criteria for DBS surgery. For inclusion into the study the following inclusion and exclusion criteria were applied: INCLUSION: Diagnosis of idiopathic Parkinson’s disease by movement disorders specialist, ^29^, meeting clinical criteria for DBS of STN/GPi, first time ever DBS surgery targeting STN or GPi bilaterally or unilaterally. EXCLUSION: Other conditions affecting gait or balance, and other neurological diseases that interfere with the purpose of the study. Figure 1 shows a flowchart of the study population with the number of patients included in the analysis at each time point.

**Figure 1.**
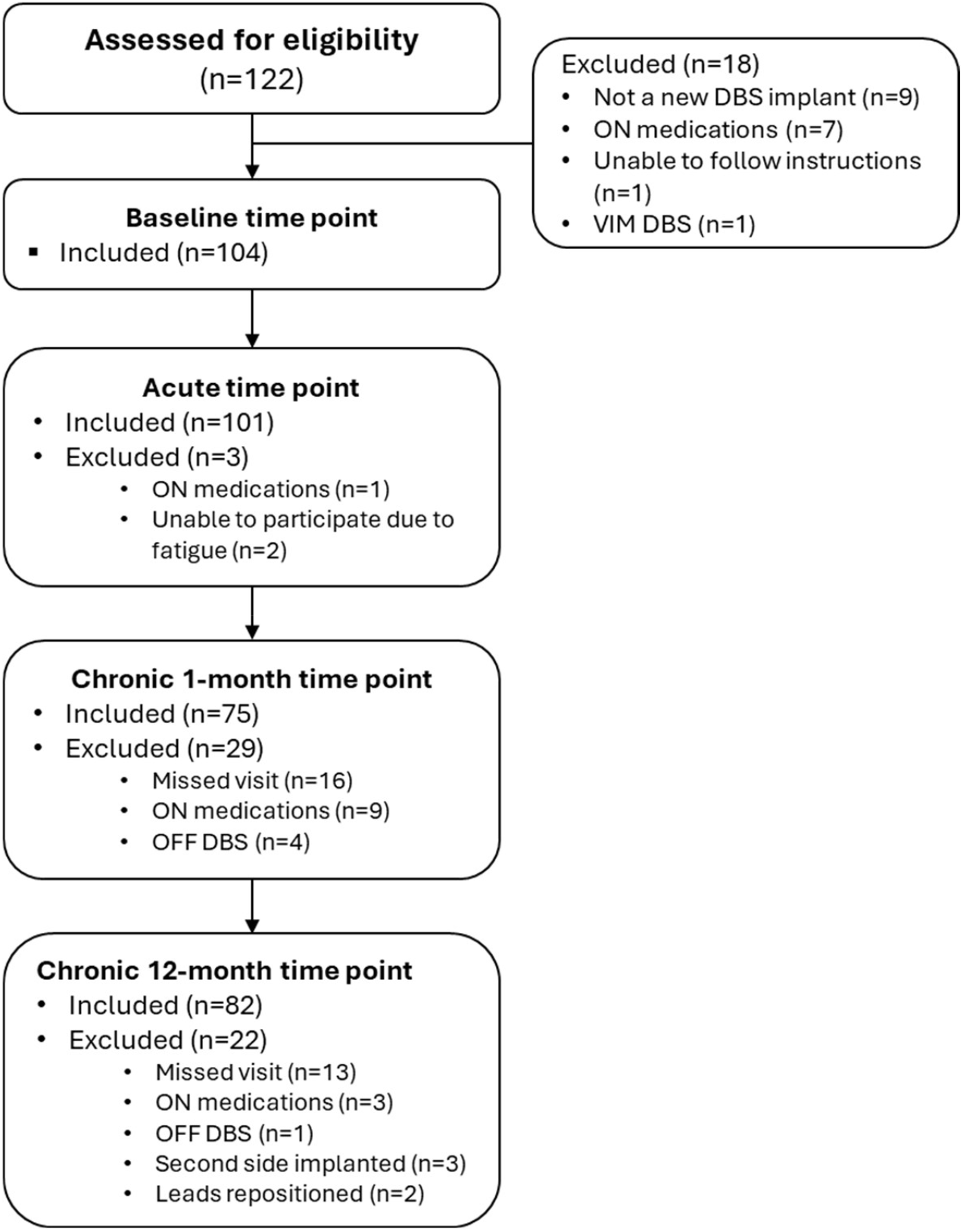
Study population: patients with Parkinson’s disease undergoing first-time STN or GPi DBS and meeting inclusion but not exclusion criteria were enrolled into the study. Data were collected at four time points: Baseline, Acute, Chronic 1-month, and Chronic 12-months.

### Standard Protocol Approvals, Registrations, and Patient Consents

The study was approved by the local Institutional Review Board and all patients signed written informed consent.

### Gait Assessments

At each visit/time point, gait kinematics were measured utilizing a 20ft. long x 4ft. wide instrumented gait mat and associated software (Protokinetics Zeno, Havertown, PA) as previously described ^30,31^. Briefly, patients traversed the walkway at their preferred speed and turned around a floor marker at the end of the walkway (alternating left/right turns). Patients performed two practice walks initially and four evaluation walks in each condition. Patients who had severe difficulty walking performed at least one walk in each condition (n=10). Patients were allowed to walk with a cane or a walker (n=7). Gait speed (cm/s) during straight walking was the primary outcome. Other gait kinematics were secondary outcomes and were divided into 5 domains as follows: pace domain including cadence (step/min), step length (cm), and stride length (cm); variability domain including swing time standard deviation (SD) (s), step time SD (s), stance time SD (s), and step length SD (cm); rhythm domain including step time (s), swing time (s), stance time (s), initial double support time (%), total double support time (%), and single support time (%); asymmetry domain including step time asymmetry (s), swing time asymmetry (s), and stance time asymmetry (s); and postural control domain including stride width (cm) and stride width SD (cm). Patients who could not walk on the walkway without assistance were defined as ‘non-ambulatory’, and their gait speed was set to zero. For individual-level analysis of gait speed, an increase or decrease of 10 cm/s was considered a clinically significant improvement or worsening, respectively ^19^. Patients who had improved/worsened gait speed at Chronic 12-months relative to Baseline were given the label “Immediate improvement/worsening” if they also improved/worsened at Acute time point and “Delayed improvement/worsening” if they only improved/worsened at 1- or 12-month time points. During each visit, patients were clinically evaluated using MDS-UPDRS part III score (motor exam), and the MDS-UPDRS axial sub-score was extracted from the MDS-UPDRS part III score using items 3.9 through 3.13 describing ability to stand from a chair, gait, freezing of gait, posture and postural stability on pull test^32^.

### Statistics

Prior to the study initiation, a power estimate for the primary analysis (gait speed (cm/s)) was done based on effect size estimates from preliminary data. At alpha=0.05, and samples of n=30 and 20, paired-sample t-tests will be able to detect moderate (7.4cm/s change) acute and chronic clinical effects of DBS with >95% and >90% power, respectively. Similar tests performed on secondary gait outcomes would have comparable power to detect moderate effect sizes.

Demographic and clinical variables were compared using analysis of variance (ANOVA) for continuous variables and chi-square tests for categorical variables. Summary statistics, including mean and SD, were calculated for gait speed and other gait kinematics. Linear mixed-effects models were used to compare gait kinematics and clinical outcomes across time points for all patients and within four DBS target groups (bilateral STN, bilateral GPi, unilateral STN, and unilateral GPi). The lmer() function from the lmerTest package (v. 3.1.3) in R (v. 4.4.1) was used. The lmerTest package extends lme4 (v. 1.1-35.5) by providing p-values for fixed effects using Satterthwaite’s approximation for degrees of freedom ^33,34^. For our primary outcome, a model for gait speed included ‘Time’ and ‘DBS target’ as fixed effects, and a random intercept for each participant. Adjusted means and change scores (mean difference) were derived from the linear mixed-effects models. Standardized effect sizes were subsequently calculated as change score divided by the standard deviation at baseline ^19,35,36^. All gait kinematic variables were then standardized using z-score normalization, and a mixed-effects linear model was utilized to calculate standardized Beta coefficient for each variable at different time points. Covariates including age, gender, disease duration, preoperative Montreal Cognitive Assessment (MoCA) scores, and preoperative levodopa equivalent daily dose (LEDD) were included to evaluate their impact on the estimated coefficients for the primary fixed effects.

For gait speed change analysis at the individual-level, we constructed a contingency table with outcome categories and DBS targets then performed a Pearson’s chi-squared test of independence using the observed frequencies and examined standardized Pearson residuals. Residuals with absolute values > 2 were considered to indicate a statistically significant departure from the expected frequency (approximate p < 0.05).

Linear regression analyses were performed to examine the relationship between clinical score changes (MDS-UPDRS part III and axial sub-score) and gait speed changes, with adjusted R^2^ values and p-values reported to assess model fit and statistical significance, respectively.

Statistical analyses were conducted using RStudio (version 2024.12.0 Build 467, Posit Software, PBC, 2009–2024). Statistical significance was set at P < 0.05. For the secondary analysis, we did not correct for multiple comparisons.

### Data Availability

Anonymized data not published within this article will be made available by request from any qualified investigator.

## RESULTS

In our study cohort of 104 patients with PD, leads were implanted bilaterally in STN (n = 45) and GPi (n = 28), or unilaterally in STN (n = 17) and GPi (n = 14) (Table 1). Baseline demographics and preoperative clinical characteristics differed significantly between the groups (Table 1). DBS target and laterality were chosen based on clinical decision making which in our center considers age, cognitive status, mood symptoms and expected tolerance for postoperative medication reduction. As a result, patients with bilateral STN DBS were the youngest (57±8 years) and patients with unilateral GPi DBS were the oldest group (71±8 years). Patients with unilateral GPi DBS had the lowest MoCA scores (24±3). Additionally, presurgical MDS-UPDRS axial sub-score OFF and ON dopaminergic medications differed significantly between groups, patients who received unilateral GPi DBS had the highest scores, followed by patients with bilateral GPi, bilateral STN, and unilateral STN, in descending order.

**Table 1.** Patient demographics and preoperative clinical characteristics: statistical analysis was performed using one-way ANOVA ^Δ^ for continuous variables and chi-square ^#^ tests for categorical variables. Values are presented as mean ± standard deviation. MoCA, Montreal Cognitive Assessment, LEDD, levodopa equivalent daily dose.

|  | Entire cohort | Bilateral STN | Unilateral STN | Bilateral GPi | Unilateral GPi | p-value |
| --- | --- | --- | --- | --- | --- | --- |
| Number of patients | 104 | 45 | 17 | 28 | 14 |  |
| Age (y) | 62±9 | 57±8 | 67±7 | 66±8 | 71±8 | <0.0001<sup>Δ</sup> |
| Gender (n) | 72 M, 32 F | 33 M, 12 F | 14 M, 3 F | 16 M, 12 F | 9 M, 5 F | 0.28<sup>#</sup> |
| Disease duration (y) | 11±5 | 10±4 | 7±3 | 12±5 | 14±5 | 0.0002<sup>Δ</sup> |
| MoCA score | 26±3 | 26±2 | 25±2 | 26±2 | 24±3 | 0.0044<sup>Δ</sup> |
| LEDD | 1399±716 | 1610±697 | 1130±592 | 1304±826 | 1239±526 | 0.055<sup>Δ</sup> |
| MDS-UPDRS I | 12.3±5 | 12.4±4.8 | 12.3±6.5 | 11.8±4.4 | 13±5.5 | 0.46<sup>Δ</sup> |
| MDS-UPDRS II | 16.3±6.8 | 15±5.5 | 15.2±5.5 | 16.4±6.6 | 21±8.6 | <0.0001<sup>Δ</sup> |
| MDS-UPDRS III – OFF medications | 44.1±11.5 | 42.8±10.6 | 42.4±12.1 | 46.7±11.4 | 45.1±13.8 | 0.018<sup>Δ</sup> |
| MDS-UPDRS III – ON medications | 16.3±8.4 | 15±9.3 | 17.9±8 | 17.1±8.1 | 16.9±7.2 | 0.051<sup>Δ</sup> |
| MDS-UPDRS axial sub-score – OFF medications | 5.6±3.4 | 5.1±2.6 | 3.9±2.7 | 6.5±4.5 | 7.4±3 | <0.0001<sup>Δ</sup> |
| MDS-UPDRS axial sub-score – ON medications | 2±1.6 | 1.7±1.3 | 1.4±1 | 2.3±2.1 | 3.2±1.3 | <0.0001<sup>Δ</sup> |
| MDS-UPDRS IV | 7.3±3.5 | 7.2±2.8 | 4.4±3.9 | 9.2±3.4 | 7.6±3 | <0.0001<sup>Δ</sup> |

At baseline, the average gait speed for the entire cohort was abnormally low at 80.6 cm/s (Fig. 2). With DBS activation, gait speed improved acutely to 97.4 cm/s (standardized effect size: 0.45, p < 0.001) and remained improved above baseline at Chronic 1-month at 93.0 cm/s (standardized effect size: 0.33, p < 0.001), and at Chronic 12-month at 91.2 cm/s (standardized effect size: 0.28, p < 0.001) (Fig. 2a, eTable 1). Covariates including age, gender, disease duration, LEDD, and MoCA score, did not significantly change the effect of DBS stimulation over time (eTable 2). When analyzed for each DBS target separately, gait speed significantly improved over baseline in the groups with bilateral STN or GPi DBS, but there was no effect with unilateral STN or GPi treatment (Fig. 2b, eTable 1). Patients with bilateral STN increased their speed from 84.7 cm/s at baseline to a Chronic 12-month gait speed of 103.6 cm/s (standardized effect size: 0.54, p< 0.001). Patients with bilateral GPi, increased their speed from 71.3 cm/s at baseline to a Chronic 12-month gait speed of 85.8 cm/s (standardized effect size: 0.33, p < 0.01) (Fig. 2b, eTable 1).

**Figure 2.**
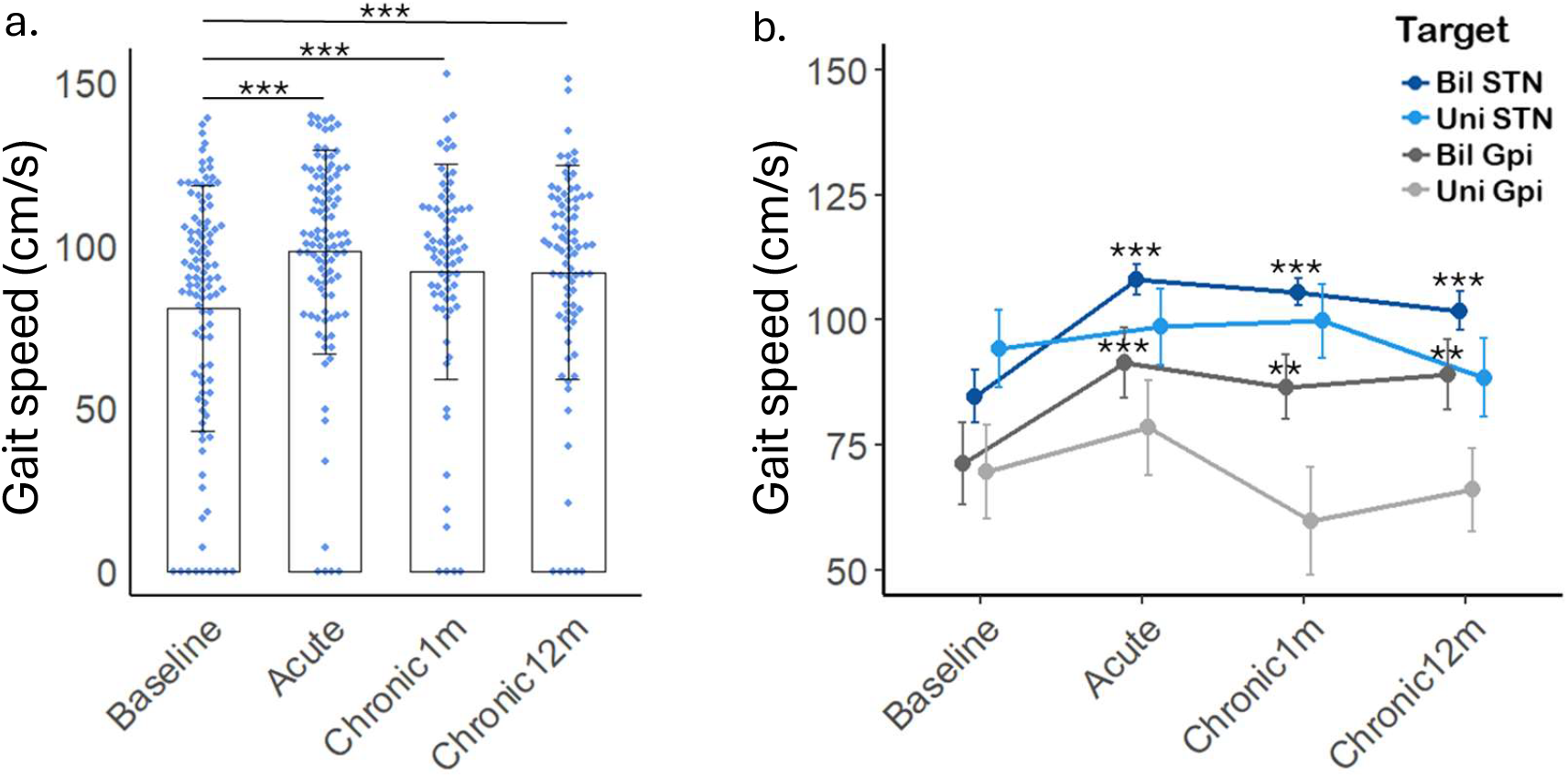
Acute and chronic effects of STN or GPi DBS on gait speed in (a) all DBS patients and (b) per DBS target group over time. Significance is represented by asterixis comparing time-points to Baseline, ^**^ p<0.01, ^***^ p<0.00. Bil, bilateral, Uni, unilateral.

Although at the group level, age did not significantly change the effect of DBS stimulation over time (eTable 2), given the age differences between target groups (Table 1) we performed a secondary analysis to explore the effect of age on gait speed in patients with bilateral STN or GPi (eFig. 1). Patients in each group were stratified by age using a median split (lower 50th percentile vs. upper 50th percentile). In patients with bilateral STN DBS, younger age (34-56 years) was related to greater increase in gait speed relative to baseline when compared to older age (57-68 years) at the Acute (30.4 vs 13.9 cm/s, p<0.05) and Chronic 12m time-points (30.5 vs 8.9 cm/s, p<0.01) (eFig. 1). This was not seen in patients with bilateral GPi (45-67 years vs 68-78 years) (eFig. 1). Note that the range of the older age group in the STN DBS group overlaps with the age range of the younger age group in the GPi DBS group.

MDS-UPDRS part III scores significantly improved after DBS activation from 44.2 ± 13.1 at Baseline to 23.1 ± 13.4 at Acute, 25.3 ± 13.0 at Chronic 1-month, and 21.2 ± 10.9 at Chronic 12-month time points (Fig 3a). Similarly, the MDS-UPDRS axial sub-score also significantly improved after DBS stimulation (Fig 3c). When analyzed by DBS target group, we found that the MDS-UPDRS part III and MDS-UPDRS axial sub-score significantly improved over time in all groups (Fig. 3b, d).

**Figure 3.**
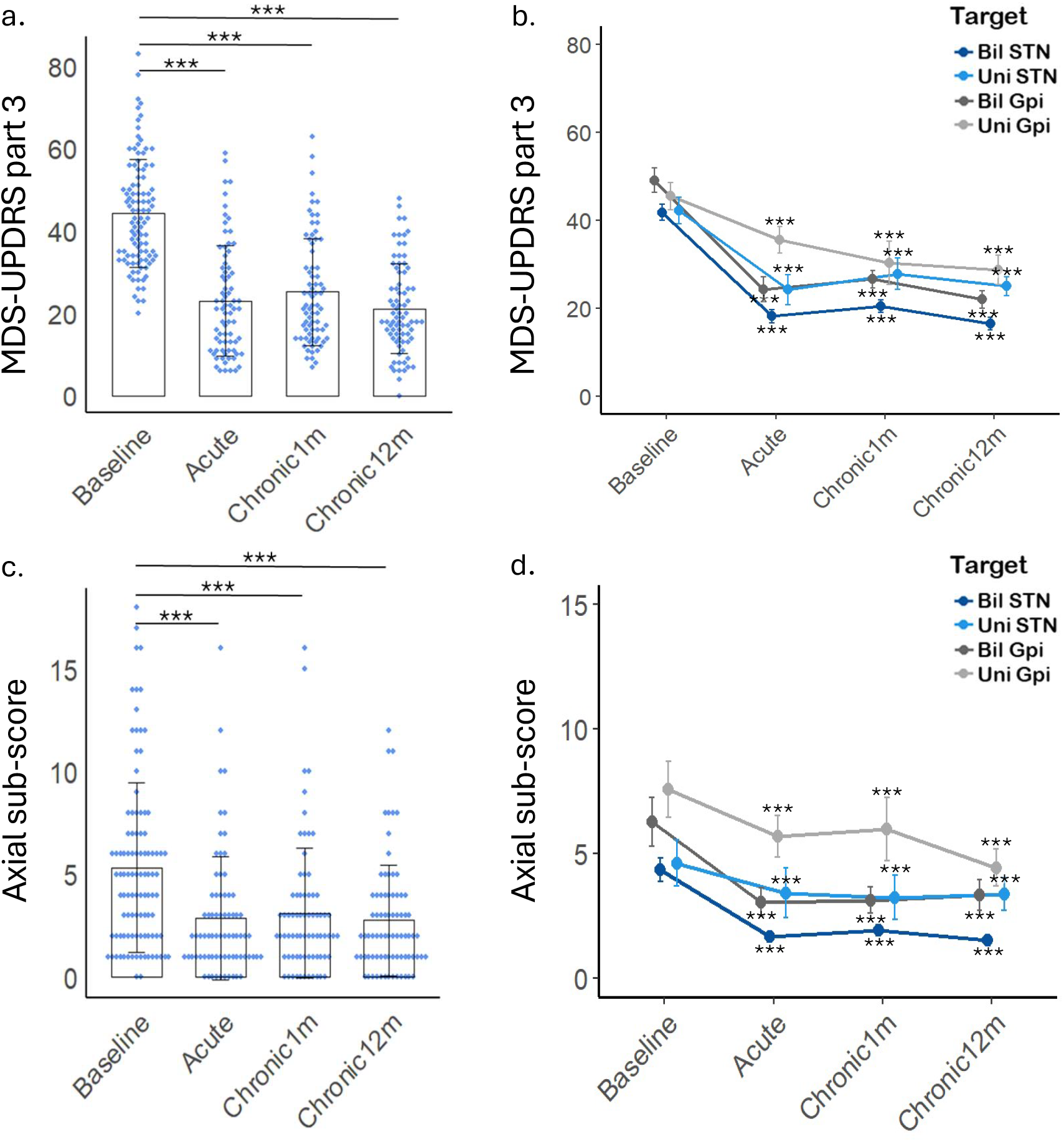
Acute and chronic effects of STN or GPi DBS on clinical scores: (a) MDS-UPDRS part III, and (c) axial sub-score in all DBS patients over time, (b) MDS-UPDRS part III, and (d) axial sub-score in DBS patients over time per DBS target group. Significance is represented by asterixis comparing time-points to Baseline, ^***^ p<0.001. Bil, bilateral, Uni, unilateral.

For more comprehensive evaluation of the effect of DBS on gait kinematics, we explored the effect of DBS therapy on kinematic gait variables belonging to 5 gait domains in all patients and by DBS target over time (Fig. 4, eFig. 2, eTable 3). Across the entire cohort, patients on average had significantly improved pace (gait speed, step length and stride length), gait variability (step time SD), and gait rhythm (swing time and single support time %) at the Acute time-point. This improvement was sustained chronically at Chronic 1-month and Chronic 12-month time-points, as compared to baseline (Fig. 4, eTable 3). Similar to the results for gait speed, only patients with bilateral STN (eFig. 2a) and bilateral GPi (eFig. 2c) had consistent improvement in gait pace, variability, and rhythm over time while patients with unilateral STN (eFig. 2b) unilateral GPi (eFig. 2d) had no significant chronic changes.

**Figure 4.**
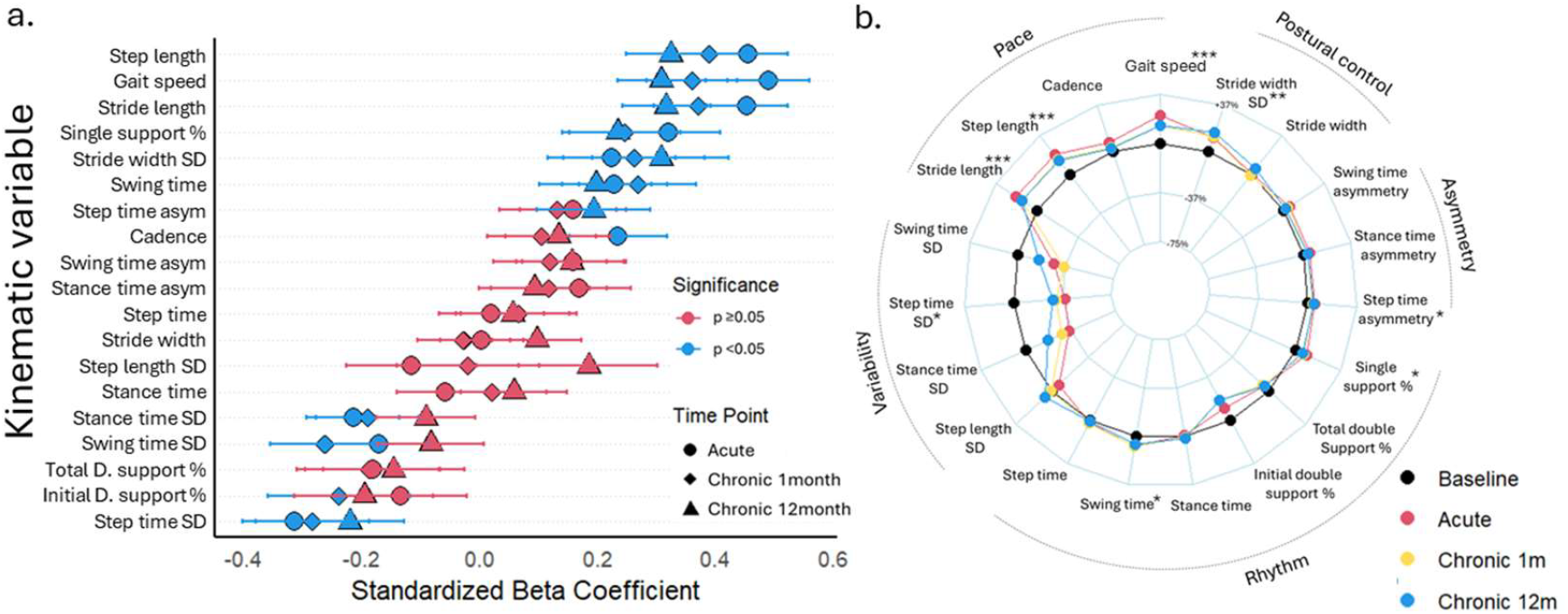
Acute and chronic effect of STN or GPi DBS on gait kinematics. (a) standardized Beta coefficient of gait variables at each time point (compared to Baseline, shape coded) with confidence interval and p value (color coded). (b) Radar plot illustrating kinematic gait outcome variables at different timepoints in all patients. The central black line represents Baseline time-point measurements, which acts as reference. Deviation from the central axis is represented as percentage change (between −75% and +37%). Significance is represented by asterixis comparing Chronic 12m to Baseline time-point, ^*^ p<0.05, ^**^ p<0.01, ^***^ p<0.00. SD, standard deviation, D., double.

To better understand the individual-level effects of DBS on gait speed over time we examined individual patient outcomes. In the entire cohort, 67/104 (64.4%) patients had impaired gait speed (less than 100 cm/s) at baseline. All patients were separated into five groups based on their direction of change (improvement vs worsening vs no change) and time to change (immediate vs delayed as detailed in Methods). At Chronic 12-month follow-up, 27% experienced a clinically meaningful improvement, 39% remained stable, and 13% worsened. Improvement was more likely to be immediate (19% vs 8%) while worsening was more likely to be delayed (10% vs 3%) (Fig. 5, eFig. 3). Distribution of outcomes differed significantly across DBS targets (χ^2^ = 25.92, df = 15, p = 0.039), and patients with bilateral STN DBS were more likely to improve immediately while those with unilateral (STN or GPi) stimulation were more likely to worsen and this often occurred immediately (Fig. 5, eFig. 3).

**Figure 5.**
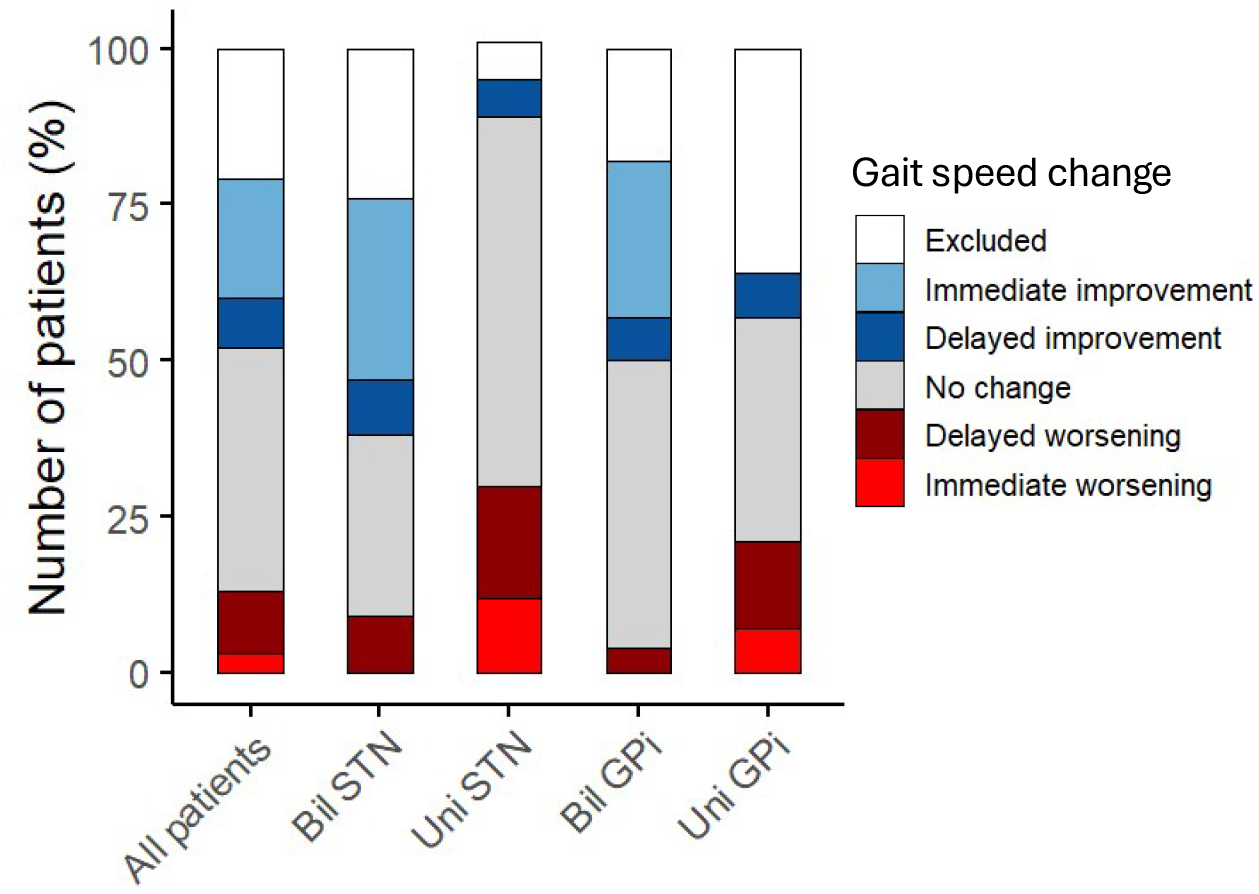
Stacked bar-graph showing the percentage of patients in five groups based on the direction of change in their gait speed (improvement vs worsening vs no change) and time to change in gait speed (immediate vs delayed).

To determine early postoperative predictors for DBS-related long-term improvement in gait, we examined the linear relationship between gait speed change at the Chronic 12-month time point and Acute and Chronic 1-month gait speed changes, and DBS-related MDS-UPDRS part III score changes. All changes were relative to Baseline gait speed and Baseline clinical scores. We found a strong positive correlation between Chronic 12-month gait speed change and both Acute (Fig. 6a, Adj. R^2^ = 0.58, p< 0.0001) and Chronic 1-month (Fig. 6b, Adj. R^2^ = 0.71, p< 0.0001) gait speed changes. This means that the direction and magnitude of gait speed change acutely and at 1-month after initial programming correlate well with gait speed outcome at 12-month follow up on the individual level. The relationship between chronic 12-month gait speed changes and clinical score changes was explored, revealing a stronger correlation with MDS-UPDRS axial sub-score changes than with MDS-UPDRS part III score changes at the acute and 1-month time points (Fig. 6 c, d, eTable 4).

**Figure 6.**
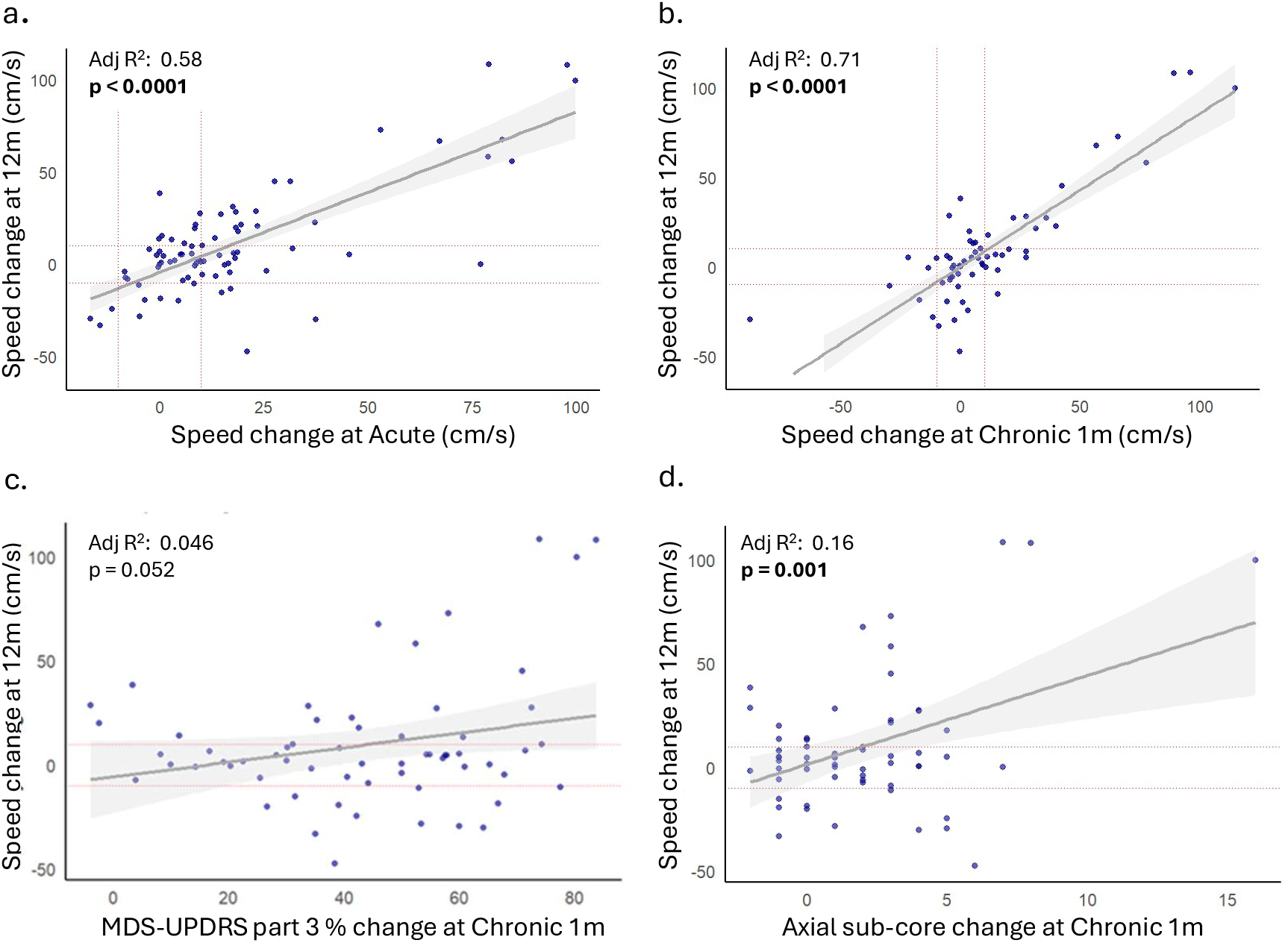
The linear relationship between speed change at (a) Acute and (b) Chronic 1-month compared to Chronic 12-month. The linear relationship between (c) MDS-UPDRS part III percent (%) change at Chronic 1-month and (d) axial sub-score change at Chronic 1-month, compared to speed change at Chronic 12-month. Speed change (Δs) is always calculated relative to baseline speed. MDS-UPDRS part III and axial sub-score Changes are always compared to baseline scores. The red dotted lines are drawn at −10cm/s and +10cm/s on the x-axis and y-axis to show clinically significant speed change thresholds. Adj, adjusted.

Given the known correlation between preoperative levodopa response on the MDS-UPDRS part III score and response to DBS ^37^, we analyzed the linear relationship between preoperative clinical levodopa response and Chronic 12-month gait speed change. In our cohort, the preoperative MDS-UPDRS part III scores improved with levodopa by at least 50% in most patients (86/104) and by at least 30% in almost all patients (99/104; data not shown). Both preoperative MDS-UPDRS part III score and preoperative axial sub-score OFF dopaminergic medications were highly correlated with gait speed at Baseline prior to DBS activation (Fig. 7a, b). However, there was no correlation between preoperative MDS-UPDRS part III levodopa response and gait speed change at Chronic 12-month (Fig. 7c, eFig. 4a, b). In contrast, the preoperative axial sub-score levodopa response did correlate (Adj R^2^: 0.05, p=0.021) with speed change at Chronic 12-month, (Fig. 7d). Secondary analysis by DBS target showed that preoperative axial sub-score levodopa response strongly correlated with speed change at Chronic 12-month only in patients with bilateral STN stimulation (Adj R^2^: 0.29, p=0.0008) and not those with bilateral GPi stimulation (Adj R^2^: 0.04, p=0. 8) (eFig. 4c, d).

**Figure 7.**
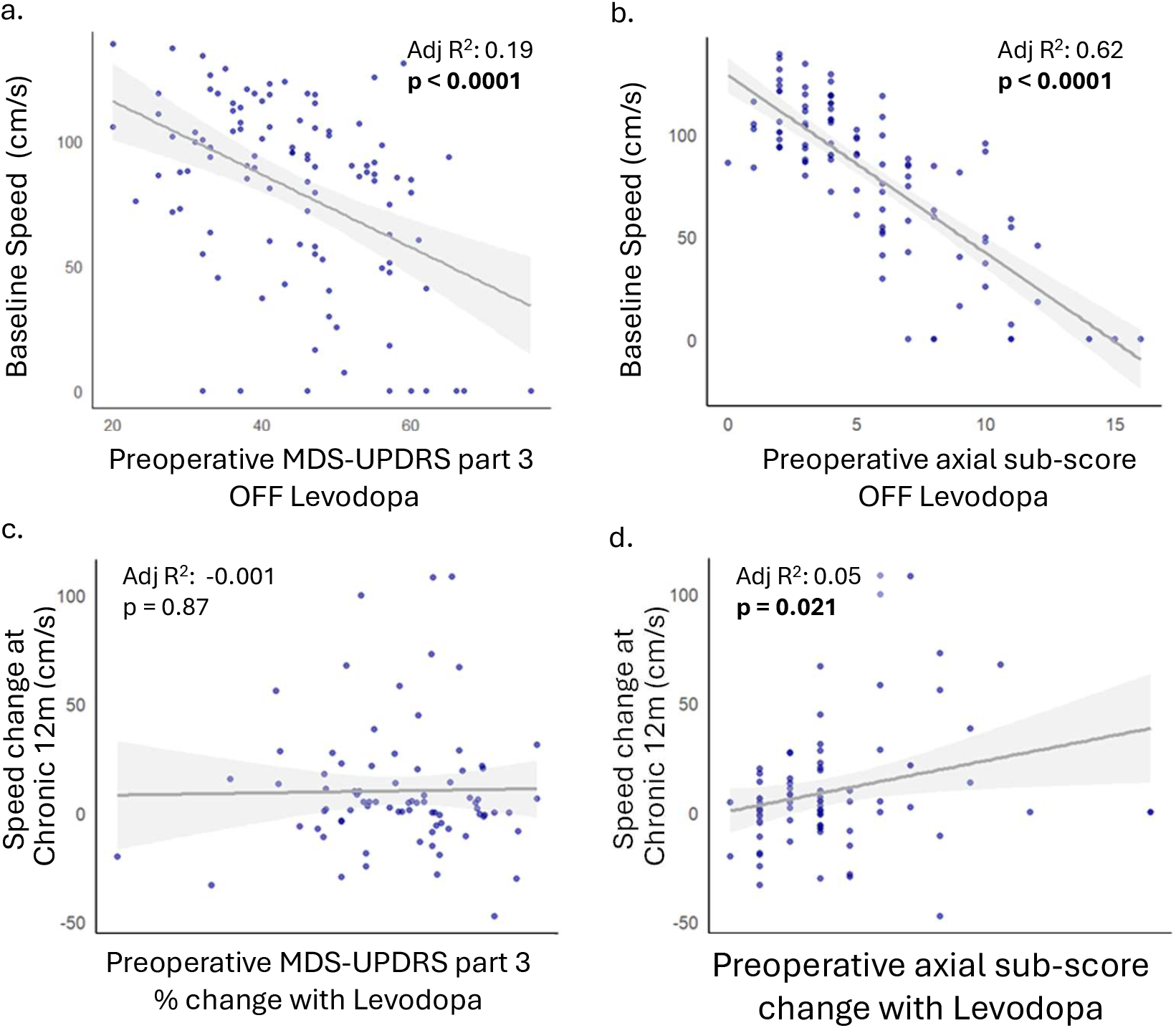
Relationship between preoperative clinical levodopa responsiveness and the effect of DBS on gait speed. (a) The linear relationship between preoperative MDS-UPDRS part III OFF levodopa score and post-DBS Baseline speed. (b) The linear relationship between preoperative axial sub-score OFF levodopa score and post-DBS Baseline speed. (c) The linear relationship between preoperative MDS-UPDRS part III % change with levodopa and speed change at Chronic 12-month. (d) The linear relationship between axial sub-score change with levodopa and speed change at Chronic 12-month. Speed change at Chronic 12-month is calculated relative to baseline speed.

## DISCUSSION

While the effects of DBS on gait have been explored since its early use, prospective, long-term studies in large PD cohorts—especially including GPi stimulation—remain sparse. By systematically studying the effect of bilateral and unilateral STN and GPi DBS in104 patients with PD longitudinally over one year in a controlled setting (ON stimulation, OFF medication), we demonstrate that gait speed significantly improved with both acute and chronic bilateral STN or GPi DBS, regardless of age, disease duration, LEDD, or MoCA score, supporting our primary hypothesis. Notably, this improvement was not observed with unilateral STN or GPi DBS treatment. Other kinematic domains of gait, including pace, variability, and rhythm, also showed significant improvement with bilateral stimulation.

Our results are similar to an earlier small cross-sectional prospective study of 18 patients with PD (8 with bilateral STN DBS, 10 with bilateral GPi DBS) ^8^. In this study, both bilateral STN and GPi stimulation were shown to improve gait motor score and gait speed and step length after 3 months of stimulation. Since this early study, the positive effect of bilateral STN DBS on gait was reported in multiple studies, some of which are well summarized in a 2016 meta-analysis of 27 studies ^6^. Here, the authors reported that bilateral STN stimulation had a moderate effect size of 0.63 on gait speed, not taking into consideration a specific time to follow up ^6^; this is similar to our finding that bilateral STN had a moderate effect size of 0.54 on gait speed after 1 year of DBS. Expanding on these findings, our study shows that bilateral GPi also exerts a significant effect on gait speed in patients with PD selected for GPi DBS therapy on clinical considerations, with a small effect size of 0.33. Because of differences in patient population when selecting between the surgical targets of GPi or STN (Table 1), the effect sizes between the groups cannot be directly compared.

In our study, unilateral STN or GPi DBS had no significant effect on gait speed at the group level and patients who worsened were more likely to receive unilateral stimulation. Worsening of gait kinematics within one year post surgery was also reported in a retrospective study of 22 patients with PD, of whom 14 were implanted with unilateral STN or GPi DBS ^38^. While direct comparison is limited due to differences in the methods, a positive effect on gait was reported when unilateral STN stimulation was turned on acutely in patients with chronic DBS ^39,40^. Our results of worsening or lack of improvement with unilateral DBS therapy could be explained by the requirement of bilateral limb activation with walking where disease progression over time can worsen gait due to contralateral limb disease involvement. Alternatively, negative effects of unilateral DBS on gait mechanics could also contribute to this result, suggested by immediate worsening in some patients (n=3) and in an earlier report by Lin et al. ^41^. The choice of unilateral versus bilateral DBS, be it STN or GPi, is guided by patient’s clinical characteristics; based on the results of our study, the presence of gait abnormalities at baseline should be considered in DBS target choice, and patients should be offered bilateral or rapidly staged DBS, when possible, to improve gait outcome.

In our cohort, gait domains of pace, variability, rhythm, and postural control showed significant improvement with bilateral STN or GPi DBS. Normative gait kinematics in patients with PD have shown that pace, variability and rhythm are the main three domains affected in patients with PD as measured both in the laboratory and free-living environments ^9,10^. Significant improvement in these domains with bilateral STN or GPi DBS would further support our hypothesis that STN and GPi DBS improve gait kinematics beyond gait speed. This can have a significant impact on quality of life in patients with PD.

In our cohort, the majority of patients who demonstrated significant improvement in gait speed at Chronic 12-months had already exhibited an Acute increase in speed shortly after intervention. Mechanistically, the improvement in gait parameters with acute stimulation suggests that it is related to immediate reduction of pathologic oscillatory neural activity with therapeutic stimulation ^22^. However, future studies should explicitly test this relationship to further support this claim. It could be helpful for the clinician to consider evaluating gait speed in addition to upper extremity symptoms at initial and early DBS programming, as those combined could better predict chronic gait changes, guiding early DBS programming for the treatment of abnormal gait. Mechanisms of long-term DBS effects on gait are still unclear but may involve its effect on plasticity in the motor cortex ^25^. The strong effect of age in patients with bilateral STN DBS, where younger age (34-56 years) was related to greater increase in gait speed when compared to older age (57-68 years) may support this type of mechanism considering the strong effect of age in plasticity of motor cortex ^42^.

Patients with a good preoperative levodopa response (>30% change on their MDS-UPDRS part III) are considered optimal DBS candidates ^26,37,43^. In our cohort, the degree of preoperative MDS-UPDRS part III levodopa responsiveness did not correlate with gait speed change post-DBS. Instead, the MDS-UPDRS axial sub-score levodopa responsiveness was predictive of post-DBS chronic speed change especially in patients with bilateral STN stimulation. In a review of literature, Fasano et al suggested that some patients with levodopa-responsive axial signs can benefit from STN or GPi DBS and noted that STN DBS might provide greater improvement of axial symptoms than GPi DBS; by contrast, GPi DBS might be associated with a milder long-term decline in axial symptoms ^4,5^. Additionally, an interesting study used the Berg balance scale (BBS) for the study on gait in patients with PD and showed that the BBS response to levodopa can predict balance improvement after bilateral STN stimulation ^44^. At this time, the MDS-UPDRS axial sub-score is an easily obtainable clinical score that can be used to predict chronic gait outcomes in patients who are planning to undergo STN DBS. Future interventional trials could evaluate the effectiveness of preoperative gait and balance scores including the axial sub-score and BBS response to levodopa in predicting long term gait outcomes in patients undergoing bilateral STN and GPi DBS.

In terms of limitations of our work, our study was not interventional, and patients were not randomized into DBS target groups. DBS target selection was based on clinical evaluation, which introduced inherent differences in patients’ baseline characteristics. However, for the group analysis, potentially confounding variables including age, disease duration, LEDD, MoCA score, and gender were accounted for by including them as co-variates into the model. The main effects were still significant after accounting for them. Patients were not blinded to their DBS on/off status which is a limitation because of a potential placebo effect. However, a placebo effect would not differ across different groups and is therefore an unlikely explanation for our findings. We did not collect pre-operative gait kinematic data for this study; hence we are unable to evaluate for any lesional effects of DBS lead placement on gait post-operatively. However, setting our baseline gait assessment 1 month post-operatively minimizes lesional effects and allows us to isolate the stimulation effect on gait by comparing our longitudinal outcomes to post-operative baseline. We did not consider freezing of gait or postural instability as outcomes in our study, mainly because we analyzed straight walking. Although some patients did have freezing of gait during straight walking, which affects gait speed, many patients with history of freezing did not freeze during straight walking.

In conclusion, bilateral STN or GPi DBS can improve gait kinematics in patients with PD. The clinician should consider the patient’s baseline gait abnormalities and MDS-UPDRS axial sub-score in DBS target and lead laterality choice and should consider evaluating gait kinematics during initial DBS programming as the response is highly predictive of long-term gait outcomes.

## Supporting information

Supplemental eFigure 1

Supplemental eFigure 2

Supplemental eFigure 3

Supplemental eFigure 4

Supplemental eTable 1

Supplemental eTable 2

Supplemental eTable 3

Supplemental eTable 4

## Data Availability

Anonymized data not published within this article will be made available to any qualified investigator by request to the authors.

## Acknowledgments

We sincerely thank the patients and their families for their participation and support. We also gratefully acknowledge the dedication and hard work of the research coordinators, whose data collection efforts were essential to the success of this study. We thank the clinical staff involved in DBS surgery and postoperative management. This work was supported by the National Institutes of Health under award number NS098685. This manuscript is the result of funding in whole or in part by the National Institutes of Health (NIH). It is subject to the NIH Public Access Policy. Through acceptance of this federal funding, NIH has been given a right to make this manuscript publicly available in PubMed Central upon the Official Date of Publication, as defined by NIH.”

## Authors’ Roles

Drs. Miocinovic, McKay, Nocera, Isbaine and Buetefisch designed the study. Drs. Al Ali, Miocinovic, Nocera, Testini, Esper, Aia, Scorr, Higginbotham, Tripathi, Au Yong, Buetefisch, along with Ms. Tran and Ms. Triche participated in patient recruitment, and data collection. Drs. Al Ali, Miocinovic, McKay and Buetefisch performed data analysis. Drs. Al Ali, Miocinovic and Buetefisch prepared the manuscript draft with important intellectual input from all other authors. All authors approved the final manuscript.

## Supplementary figure legends and table titles

**eTable 1:** Average gait speed changes at Acute, Chronic 1-month, and Chronic 12-month, as compared to baseline average speed (cm/s) in all patients and per DBS target groups, reported as change score (cm/s) and standardized effect size with 95% confidence intervals. Std, standardized, SD, standard deviation, n, sample size.

**eTable 2:** Main mixed-effects model Beta coefficients for gait speed and it’s percent change with the addition of covariates to the main mixed-effects model

**eTable 3:** Kinematic gait outcome variables over time in all patients, grouped across five gait domains. Values are presented as mean (standard deviation). Statistical significance is compared to Baseline, ^*^ p<0.05, ^**^ p<0.01, ^***^ p<0.001

**eTable 4:** Linear model results of Acute and Chronic 1-month speed changes and clinical score changes correlated with chronic 12-month speed change (Δs). Clinical scores included MDS-UPDRS part 3 and axial sub-score and were calculated as arithmetic change or percent (%) change. All changes were compared to Baseline.

**eFigure 1**. Effect of age on bilateral STN and GPi gait speed over time in patients with (a) bilateral STN and (b) bilateral GPi, stratified by age using a median split (lower 50th percentile vs. upper 50th percentile), with the age range depicted for reference. ^*^ p<0.05, ^**^ p<0.01

**eFigure 2**. Radar plot illustrating kinematic gait outcome variables at different timepoints in patients with (a) bilateral STN DBS, (b) unilateral STN DBS, (c) bilateral GPi DBS, and (d) unilateral GPi DBS. The central black line represents Baseline time-point measurements, which acts as reference. Deviation from the central axis is represented as percentage change (between −75% and +37%). Significance is represented by asterixis comparing Chronic 12m to Baseline time-point, ^*^ p<0.05, ^**^ p<0.01, ^***^ p<0.00. SD, standard deviation, D., double.

**eFigure 3**. (a) Contingency table showing the number of patients per gait speed change outcome category (DI, DW, II, IW, NC, or U) for each DBS target. Distribution of outcomes differed significantly across DBS targets (χ^2^ = 25.92, df = 15, p = 0.039). (b) Heatmap of standardized Pearson residuals from the chi-squared test of independence between DBS target and clinical outcome categories. Positive residuals (red) indicate more observations than expected, and negative residuals (blue) indicate fewer than expected. Cells with absolute residuals > 2 are considered to significantly contribute to the overall chi-squared statistic (approximately p < 0.05). DI, delayed improvement, DW, delayed worsening, II, immediate improvement, IW, immediate worsening, NC, no change, U, unknown.

**eFigure 4**. Linear relationship between preoperative MDS-UPDRS part III % change with levodopa and speed change at Chronic 12-month in patients with (a) bilateral STN and (b) bilateral GPi. Linear relationship between preoperative axial sub-score change with levodopa and speed change at Chronic 12-month in patients with (c) bilateral STN and (d) bilateral GPi. Speed change at Chronic 12-month is calculated relative to Baseline speed.

