## Supplemental eFigure 1 for "Bilateral Deep Brain Stimulation of the Subthalamic Nucleus or Globus Pallidus Internus Improves Gait Impairment in Parkinson’s Disease"

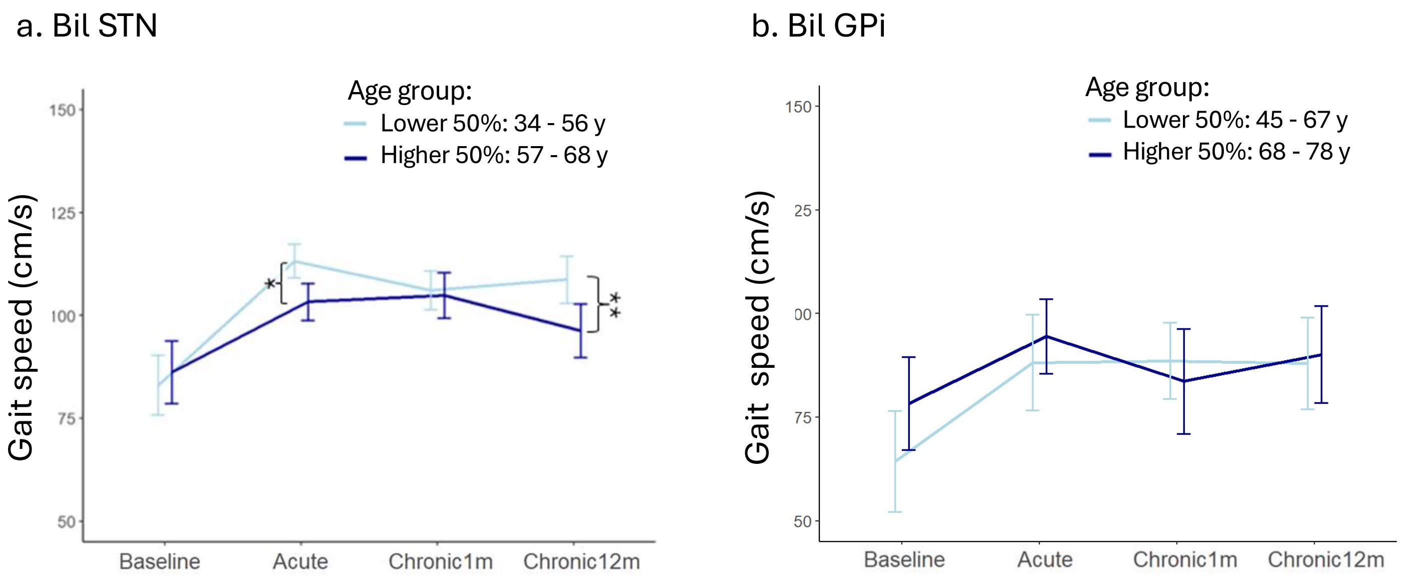


**eFigure 1**. Effect of age on bilateral STN and Gpi gait speed over time in patients with (a) bilateral STN and (b) bilateral GPi, stratified by age using a median split (lower 50th percentile vs. upper 50th percentile), with the age range depicted for reference. * $p<0.05$, ** $p<0.01$
