## Supplemental eFigure 2 for "Bilateral Deep Brain Stimulation of the Subthalamic Nucleus or Globus Pallidus Internus Improves Gait Impairment in Parkinson’s Disease"

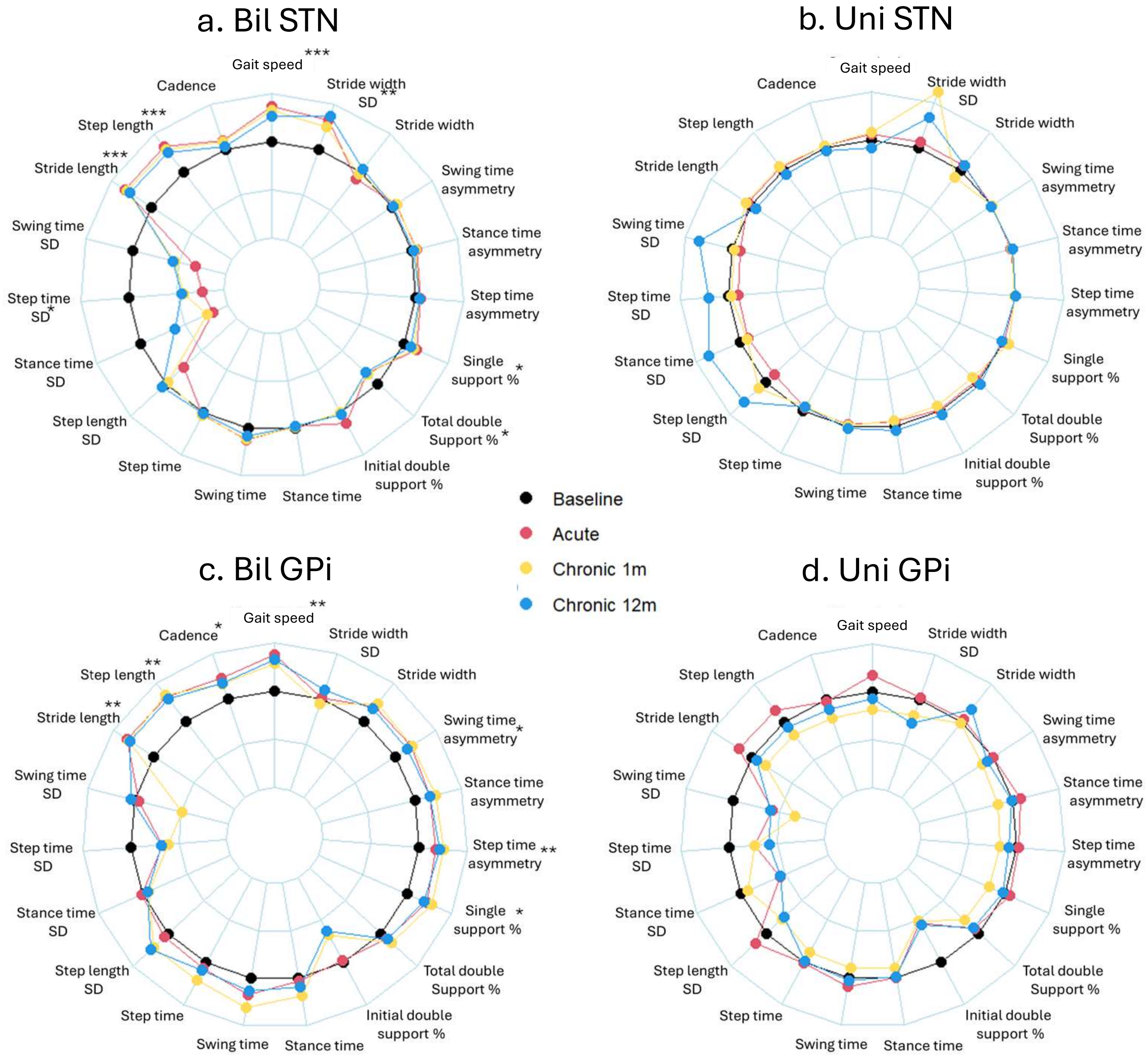


**eFigure 2:** Radar plot illustrating kinematic gait outcome variables at different timepoints in patients with (a) bilateral STN DBS, (b) unilateral STN DBS, (c) bilateral GPi DBS, and (d) unilateral GPi DBS . The central black line represents Baseline time-point measurements, which acts as reference. Deviation from the central axis is represented as percentage change (between -75% and +37%). Significance is represented by asterixis comparing Chronic 12m to Baseline time-point, * p<0.05, ** p<0.01, *** p<0.00. SD, standard deviation, D., double.
