## Supplemental eFigure 3 for "Bilateral Deep Brain Stimulation of the Subthalamic Nucleus or Globus Pallidus Internus Improves Gait Impairment in Parkinson’s Disease"

a.

| | DI | DW | II | IW | NC | U |
|---|---|---|---|---|---|---|
| Bilateral GPi | 2 | 1 | 7 | 0 | 13 | 5 |
| Bilateral STN | 4 | 4 | 13 | 0 | 13 | 11 |
| Unilateral GPi | 1 | 2 | 0 | 1 | 5 | 5 |
| Unilateral STN | 1 | 3 | 0 | 2 | 10 | 1 |

b.

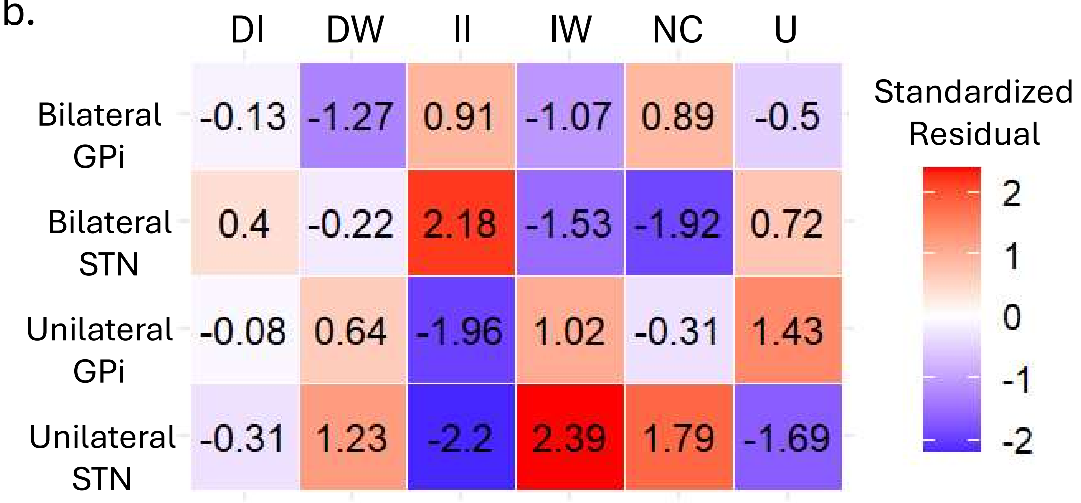


**eFigure 3**. (a) Contingency table showing the number of patients per gait speed change outcome category (DI, DW, II, IW, NC, or U) for each DBS target. Distribution of outcomes differed significantly across DBS targets ($\chi^2 = 25.92$, df = 15, $p = 0.039$). (b) Heatmap of standardized Pearson residuals from the chi-squared test of independence between DBS target and clinical outcome categories. Positive residuals (red) indicate more observations than expected, and negative residuals (blue) indicate fewer than expected. Cells with absolute residuals > 2 are considered to significantly contribute to the overall chi-squared statistic (approximately $p < 0.05$). DI, delayed improvement, DW, delayed worsening, II, immediate improvement, IW, immediate worsening, NC, no change, U, unknown.
