## Supplemental eFigure 4 for "Bilateral Deep Brain Stimulation of the Subthalamic Nucleus or Globus Pallidus Internus Improves Gait Impairment in Parkinson’s Disease"

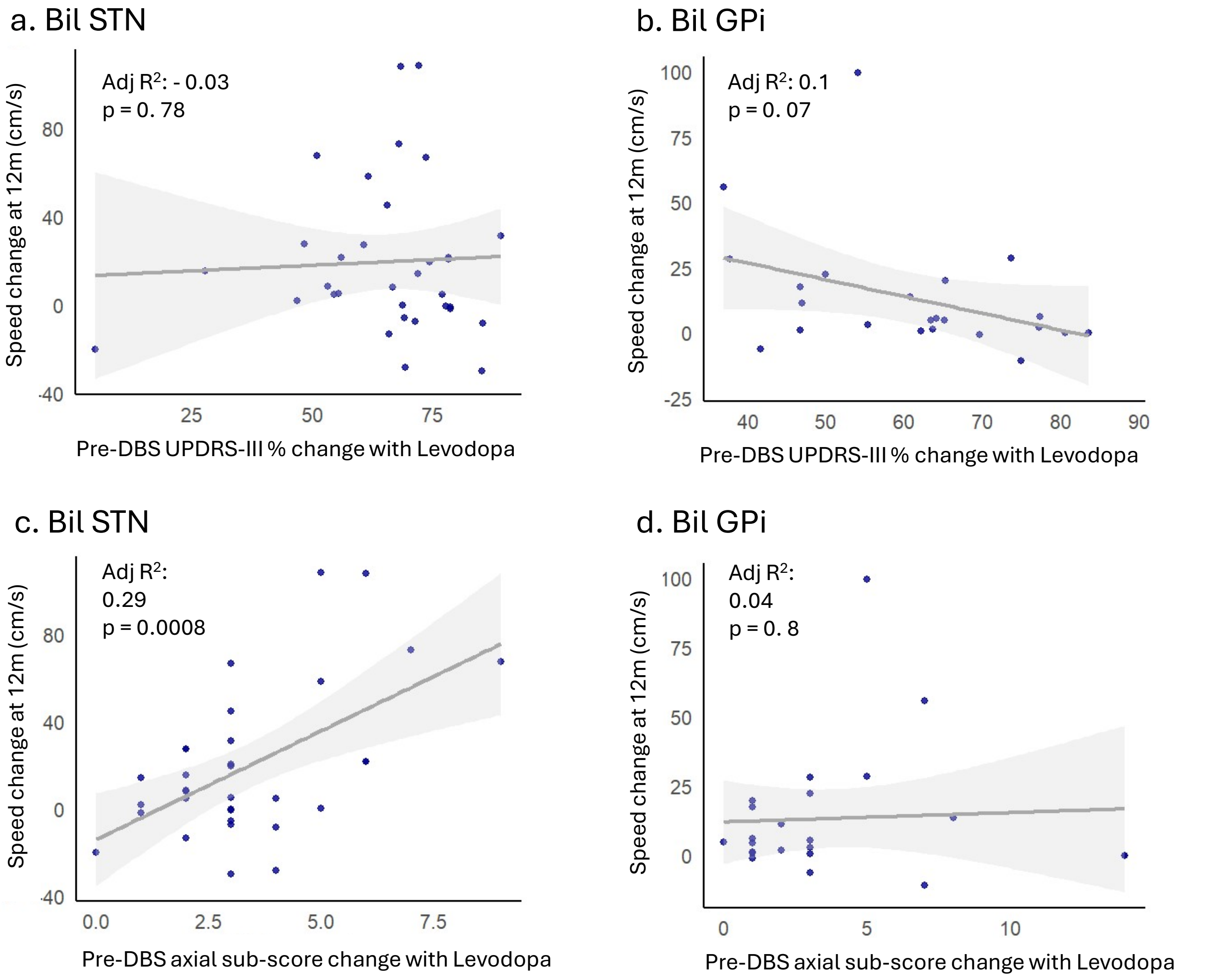


**eFigure 4**. Linear relationship between pre-DBS MDS-UPDRS-III % change with levodopa and speed change at Chronic 12-month in patients with (a) bilateral STN and (b) bilateral Gpi. Linear relationship between pre-DBS axial sub-score change with levodopa and speed change at Chronic 12-month in patients with (c) bilateral STN and (d) bilateral Gpi. Speed change at Chronic 12-month is calculated relative to Baseline speed
