## Supplemental eTable 1 for "Bilateral Deep Brain Stimulation of the Subthalamic Nucleus or Globus Pallidus Internus Improves Gait Impairment in Parkinson’s Disease"

**eTable 1**: Average gait speed changes at Acute, Chronic 1-month, and Chronic 12-month, as compared to baseline average speed (cm/s) in all patients and per DBS target groups, reported as change score (cm/s) and standardized effect size with 95% confidence intervals. Std, standardized, SD, standard deviation, n, sample size.

| | Baseline | | | Acute | | | Chronic 1-month | | | Chronic 12-month | | |
|---|---|---|---|---|---|---|---|---|---|---|---|---|
| | Baseline speed | n | SD | Change score | n | Std effect size | Change score | n | Std effect size | Change score | n | Std effect size |
| All patients | 80.6 [73.98, 87.23] | 104 | 37.5 | 16.8 [11.12, 22.48] | 101 | 0.45 [0.24, 0.65] | 12.35 [7.15, 17.56] | 75 | 0.33 [0.10, 0.56] | 10.58 [5.55, 15.62] | 82 | 0.28 [0.06, 0.50] |
| Bilateral STN | 84.7 [74.84, 94.56] | 45 | 35.0 | 21.8 [15.91, 27.69] | 43 | 0.62 [0.30, 0.95] | 21 [12.53, 29.47] | 26 | 0.60 [0.18, 1.02] | 18.9 [11.16, 26.64] | 34 | 0.54 [0.18, 0.90] |
| Unilateral STN | 94.2 [77.43, 111.03] | 17 | 31.6 | 4.34 [−6.77, 15.45] | 17 | 0.14 [-0.34, 0.61] | 3.69 [−7.62, 15.00] | 16 | 0.12 [-0.38, 0.61] | -5.73 [−17.05, 5.60] | 16 | -0.18 [-0.68, 0.31] |
| Bilateral GPi | 71.3 [58.67, 84.00] | 28 | 43.5 | 20.38 [11.67, 29.10] | 27 | 0.47 [0.07, 0.87] | 14.39 [4.95, 23.82] | 22 | 0.33 [-0.10, 0.76] | 14.48 [5.15, 23.81] | 23 | 0.33 [-0.09, 0.75] |
| Unilateral GPi | 69.6 [50.71, 88.42] | 14 | 35.1 | 8.9 [-3.37, 21.17] | 14 | 0.25 [-0.28, 0.79] | -4.5 [-17.89, 8.89] | 11 | -0.13 [-0.72, 0.47] | -2.9 [-17.29, 11.50] | 9 | -0.08 [-0.74, 0.57] |
