## Supplemental eTable 2 for "Bilateral Deep Brain Stimulation of the Subthalamic Nucleus or Globus Pallidus Internus Improves Gait Impairment in Parkinson’s Disease"

**eTable 2** : Main mixed-effects model Beta coefficients for gait speed and it's percent change with the addition of covariates to the main mixed-effects model

| | Acute | Chronic 1m | Chronic 12m |
|---|---|---|---|
| Main model β coefficient for gait speed (cm/s) | 16.8 | 12.4 | 10.6 |
| β coefficient change with age (%) | 0.1 | 0.1 | 0.0 |
| β coefficient change with disease duration (%) | 0.2 | -0.3 | -0.7 |
| β coefficient change with LEDD (%) | 2.9 | 11.7 | 4.8 |
| β coefficient change with MoCA (%) | 2.9 | 11.8 | 6.4 |
| β coefficient change with Gender: Male (%) | 0.7 | 30.4 | -3.5 |
