## Supplemental eTable 3 for "Bilateral Deep Brain Stimulation of the Subthalamic Nucleus or Globus Pallidus Internus Improves Gait Impairment in Parkinson’s Disease"

**eTable 3** : Kinematic gait outcome variables over time in all patients, grouped across five gait domains. Values are presented as mean (standard deviation). Statistical significance is compared to Baseline, * p<0.05, ** p<0.01, *** p<0.001

| Gait Domain | Gait Variable | Baseline | Acute | Chronic 1-month | Chronic 12-months |
|---|---|---|---|---|---|
| **Pace** | Gait speed (cm/s) | 80.6 (37.5) | 97.9 (31.1) *** | 92 (33) *** | 91.7 (33) *** |
| | Cadence (step/min) | 94.9 (32.2) | 101.7 (22.7) ** | 97.7 (25.9) | 97.5 (27.6) |
| | Step length (cm) | 46.2 (21) | 55.1 (16.8) *** | 53.1 (17.3) *** | 52.3 (17.8) *** |
| | Stride length (cm) | 92.9 (41.9) | 110.7 (33.8) *** | 105.7 (35.4) *** | 105.4 (35.7) *** |
| **Variability** | Swing time SD (s) | 0.04 (0.08) | 0.03 (0.07) * | 0.02 (0.02) ** | 0.03 (0.07) |
| | Step time SD (s) | 0.05 (0.09) | 0.03 (0.04) *** | 0.03 (0.03) ** | 0.03 (0.04) * |
| | Stance time SD (s) | 0.07 (0.13) | 0.04 (0.11) ** | 0.05 (0.09) * | 0.06 (0.13) |
| | Step length SD (cm) | 3.43(1.9) | 3.2 (1.7) | 3.5 (2.4) | 3.7 (2.3) |
| **Rhythm** | Step time (s) | 0.54 (0.2) | 0.55 (0.1) | 0.55 (0.2) | 0.55 (0.2) |
| | Swing time (s) | 0.34 (0.1) | 0.37 (0.1) * | 0.37 (0.1) ** | 0.36 (0.1) * |
| | Stance time (s) | 0.74 (0.3) | 0.73 (0.2) | 0.75 (0.2) | 0.75 (0.3) |
| | Initial double support time (%) | 18.9 (24.5) | 17 (17.5) | 15.5 (5.3) * | 15.6 (5) |
| | Total double support time (%) | 32.1 (16.9) | 30 (8.2) | 30.1 (8.5) | 31 (9) |
| | Single support time (%) | 29.7 (10.2) | 32.5 (7.7) *** | 32 (8.4) ** | 31.4 (8.7) * |
| **Asymmetry** | Step time asymmetry (s) | 0.92 (0.3) | 0.97 (0.21) | 0.96 (0.24) | 0.97 (0.26) * |
| | Swing time asymmetry (s) | 0.92 (0.32) | 0.97 (0.21) | 0.95 (0.24) | 0.96 (0.26) |
| | Stance time asymmetry (s) | 0.91 (0.29) | 0.96 (0.2) | 0.95 (0.23) | 0.93 (0.24) |
| **Postural Control** | Stride width (cm) | 6.9 (4.3) | 7 (4.1) | 6.9 (4.2) | 7.4 (4) |
| | Stride width SD (cm) | 1.9 (1) | 2.1 (1) * | 2.1 (1.1) * | 2.2 (0.9) ** |
