## Supplemental eTable 4 for "Bilateral Deep Brain Stimulation of the Subthalamic Nucleus or Globus Pallidus Internus Improves Gait Impairment in Parkinson’s Disease"

**eTable 4:** Linear model results of Acute and Chronic 1-month speed changes and clinical score changes correlated with chronic 12-month speed change (Δs). Clinical scores included MDS-UPDRS part 3 and axial sub-score and were calculated as arithmetic change or percent (%) change. All changes were compared to Baseline.

| Linear model | Adj R2 | p-value |
|---|---|---|
| Chronic 12m Δs vs Acute Δs | 0.58 | <0.0001 |
| Chronic 12m Δs vs Chronic 1m Δs | 0.71 | <0.0001 |
| Chronic 12m Δs vs Acute MDS-UPDRS part 3 change | 0.032 | 0.094 |
| Chronic 12m Δs vs Acute MDS-UPDRS part 3 % change | 0.039 | 0.072 |
| Chronic 12m Δs vs Chronic 1m MDS-UPDRS part 3 change | 0.079 | 0.015 |
| Chronic 12m Δs vs Chronic 1m MDS-UPDRS part 3 % change | 0.046 | 0.052 |
| Chronic 12m Δs vs Acute axial change | 0.071 | 0.024 |
| Chronic 12m Δs vs Acute axial % change | 0.026 | 0.118 |
| Chronic 12m Δs vs Chronic 1m axial change | 0.160 | 0.001 |
| Chronic 12m Δs vs Chronic 1m axial % change | 0.021 | 0.139 |
